# More than medications: A patient-centered assessment of Parkinson’s disease care needs during hospitalization

**DOI:** 10.1101/2023.07.18.23292580

**Authors:** Jessica Shurer, Shannon L. S. Golden, Paul Mihas, Nina Browner

**Affiliations:** CurePSP, Inc., New York, NY; Goldsmith Research Group, Winston-Salem, NC; Odum Institute for Research in Social Science, University of North Carolina at Chapel Hill; Department of Neurology, University of North Carolina at Chapel Hill

**Keywords:** Parkinson’s disease, Hospitalization, Patient-centered care, qualitative methods, focus groups

## Abstract

**Background:** Parkinson’s disease (PD) increases the risk of hospitalization and complications while in the hospital. Patient-centered care emphasizes active participation of patients in decision-making and has been found to improve satisfaction with care. Engaging in discussion and capturing hospitalization experience of a person with PD (PwP) and their family care partner (CP) is a critical step towards the development of quality improvement initiatives tailored to the unique hospitalization needs of PD population.

**Objectives:** This qualitative study aimed to identify the challenges and opportunities for PD patient-centered care in hospital setting.

**Methods:** Focus groups were held with PwPs and CPs to capture first-hand perspectives and generate consensus themes on PD care during hospitalization. A semi-structured guide for focus group discussions included questions about inpatient experiences and interactions with the health system and clinical team. Data was analyzed using inductive thematic analysis.

**Results:** A total of twelve PwPs and thirteen CPs participated in seven focus groups. Participants were 52% female and 28% nonwhite; 84% discussed unplanned hospitalizations. This paper focuses on two specific categories that emerged from the data analysis. The first category explores the impact of PD diagnosis on the hospital experience, specifically during planned and unplanned hospitalizations. The second category delves into the unique needs of PwPs and CPs during hospitalization, which included the importance of proper PD medication management, the need for improved hospital ambulation protocols, and the creation of disability informed hospital environment specific for PD.

**Conclusion:** PD diagnosis impacts the care experience, regardless of the reason for hospitalization. While provision of PD medications was a challenge during hospitalization, participants also desired flexibility of ambulation protocols and an environment that accommodated their disability. Findings highlight the importance of integrating the perspectives of PwPs and CPs when targeting patient-centered interventions to improve hospital experiences and outcomes.

---

People with a clinical diagnosis of Parkinson’s disease (PD) experience more frequent and prolonged hospitalizations than their age-matched peers.^1–6^ Most hospitalizations occur on general wards and result from a comorbid disorder or health crisis such as respiratory and urinary tract infections, cardiovascular diseases, falls and fractures. ^7–12^ It is well documented that during hospitalizations, person with PD (PwP) is at a higher risk of complications, including falls, medication errors, development of delirium and psychosis, and overall decline of their pre- existing motor and non-motor symptoms of PD. ^8,13–16^ Improved medication adherence during hospital stays, e-alerts to PD specialists upon admission, development of the Parkinson’s Foundation Aware in Care hospital kit, and recommendations for ward certification programs are among the calls to action and quality improvement interventions that have targeted hospital outcomes for PD. ^17–22^ However, to date, only a few have resulted in significant decreases in the length of stay or complications during hospitalization for PD. ^18,23^

In the context of inpatient hospitalization for older adults and those with chronic and serious medical conditions, patient-centered care (PCC) has revealed benefits in intermediate and distal outcomes, and almost all studies found positive relationships between PCC approaches and patient satisfaction. ^24–27^ PCC places patients at the center of the healthcare decision-making process and recognizes the importance of their individual preferences and goals. ^28,29^ Growing awareness of PCC delivery resulted in the establishment of specialized multidisciplinary teams as the gold standard of outpatient care for PD as well as the increasing application of palliative care, a traditionally team-based model of care, for the management of the physical, emotional, and spiritual needs of PwPs. ^30–36^

Presently, the voices of PwPs and their care partners (CPs) regarding their experiences during hospitalization are not well represented in the literature, and most research on PCC for PD has focused on outpatient care. Studies that qualitatively investigated hospitalization for PD primarily highlighted medication mismanagement, struggles with postoperative confusion, and deterioration of motor symptoms; however, they minimally captured patient-reported needs or experiences of CPs during hospitalization. ^37–41^ To bridge this gap and to expand the scope of patient-centered PD care from outpatient to inpatient settings, our study gathered and analyzed valuable self-reported experiences of PwPs and CPs regarding hospitalization.

## Methods

### Study design

Focus groups were selected as the optimal methodology to capture first-hand experiences of PwPs and CPs and to generate themes on PD care during hospitalization. ^42,43^ Participants either had a neurologist-confirmed clinical diagnosis of PD or were a family member of a person with a neurologist-confirmed clinical diagnosis of PD and were able to participate in the interview. Hospitalization was defined as a planned (e.g., scheduled surgery or procedure) or unplanned (e.g., emergent or urgent admission) hospital stay for at least 24 hours between January 2018 and July 2022. Patients hospitalized for deep brain stimulation surgery were excluded. All participants had to be at least 18 years of age, with no upper age limit.

This study was conducted in accordance with the Declaration of Helsinki and was approved and overseen by the University of North Carolina at Chapel Hill Institutional Review Board. Participants provided verbal informed consent prior to any study activity, including data collection. This study was supported by a Parkinson’s Foundation Community Outreach Resource Education grant.

### Data Collection

Participants were recruited through clinician referrals, announcements shared with North Carolina-based PD support groups, and flyers placed at the outpatient neurology clinic at the University of North Carolina at Chapel Hill. A purposive sampling strategy was used to recruit a variety of PwPs and CPs with a range of hospital experiences (Table 1). Quotas were applied to the patient sample to ensure a diverse representation, including demographic (e.g., sex, age, race) and clinical characteristics (e.g., stage of condition as defined by Hoehn and Yahr score, planned versus unplanned hospitalization experiences). ^44^ Half-way through recruitment, additional efforts were made to enroll participants from underrepresented demographics of the study.

**Table 1.**
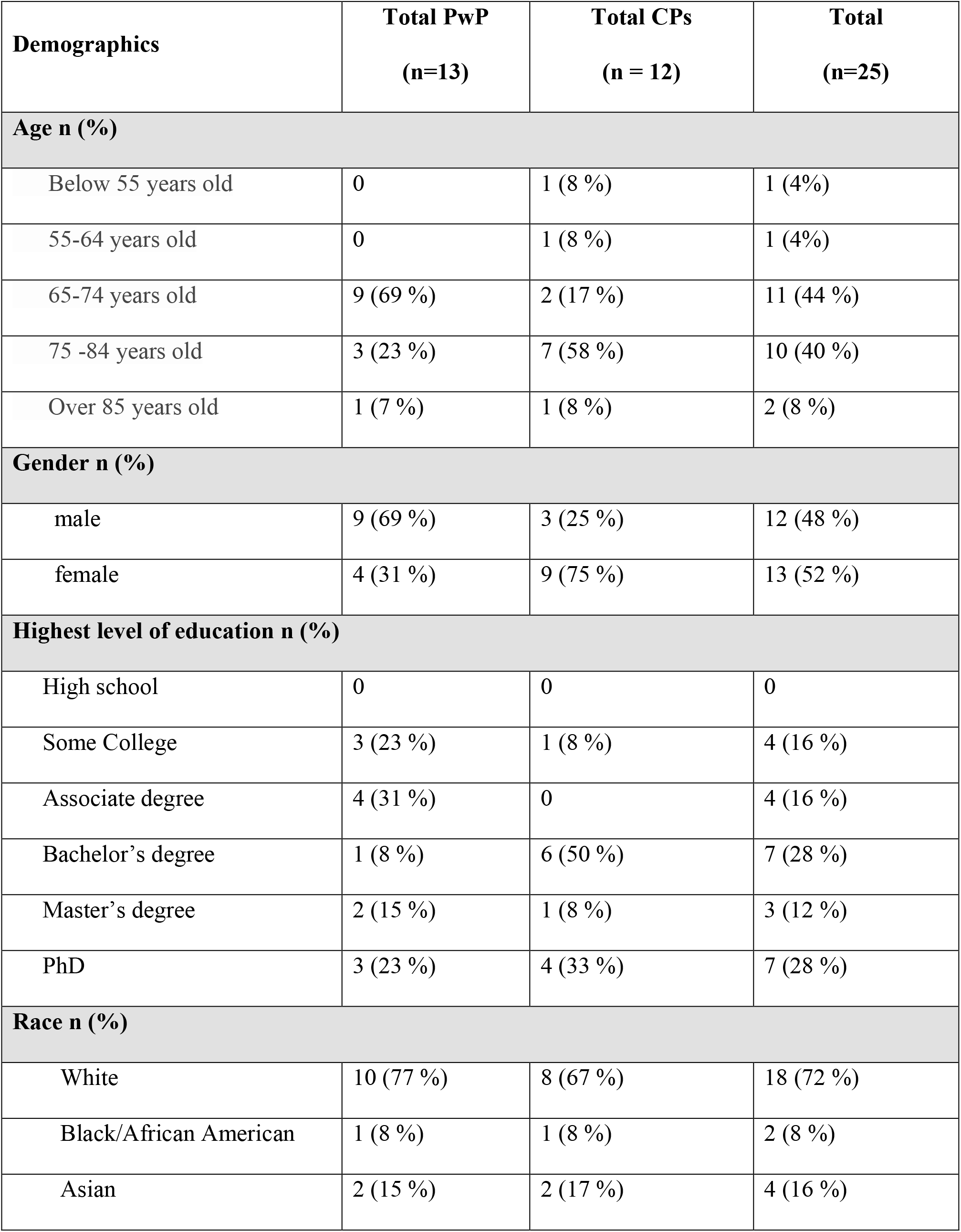
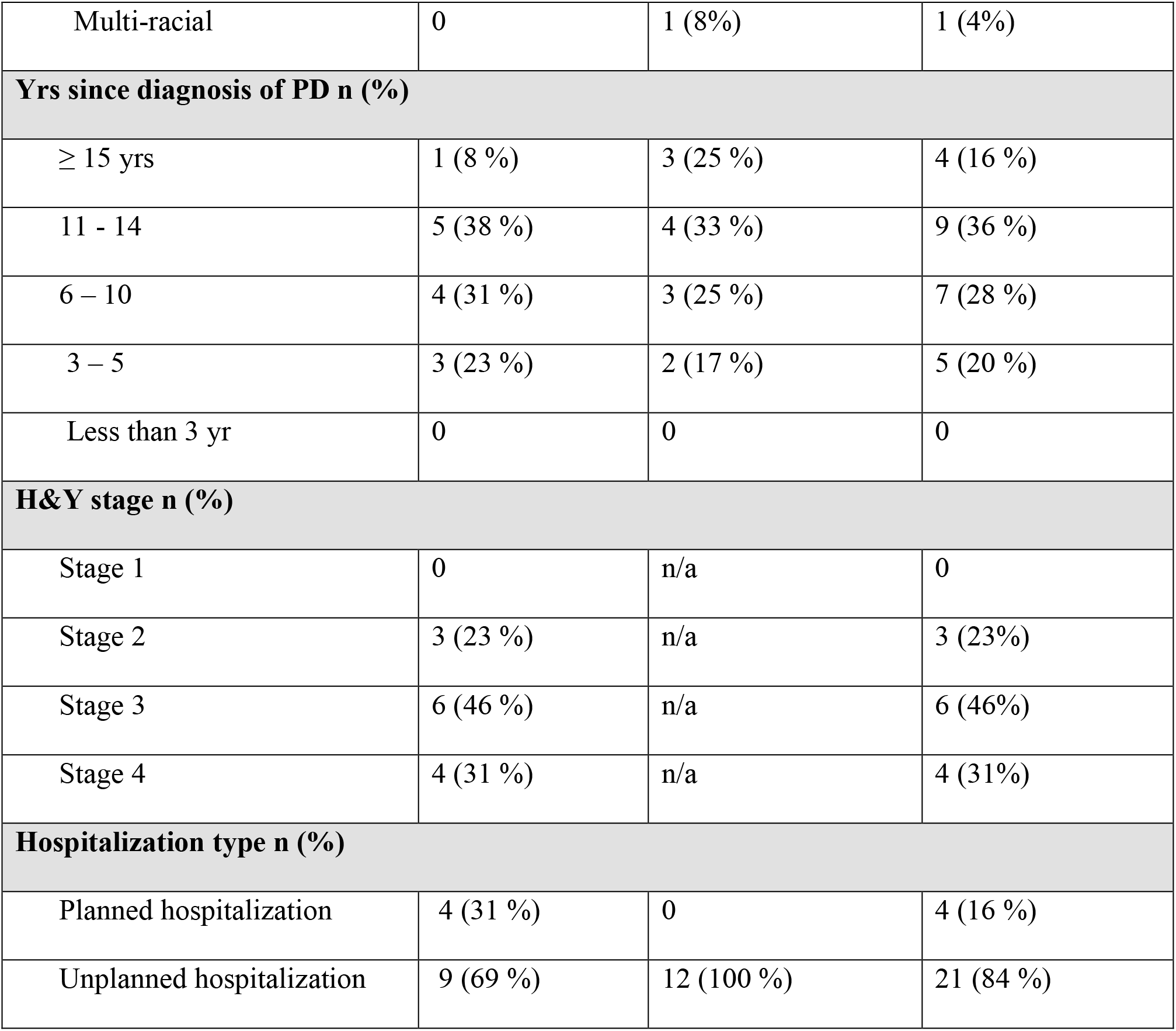
Demographic and clinical characteristics of the participants.

| **Demographics** | **Total PwP**<br>**(n=13)** | **Total CPs**<br>**(n = 12)** | **Total**<br>**(n=25)** |
| --- | --- | --- | --- |
| **Age n (%)** |  |  |  |
| Below 55 years old | 0 | 1 (8 %) | 1 (4%) |
| 55-64 years old | 0 | 1 (8 %) | 1 (4%) |
| 65-74 years old | 9 (69 %) | 2 (17 %) | 11 (44 %) |
| 75 -84 years old | 3 (23 %) | 7 (58 %) | 10 (40 %) |
| Over 85 years old | 1 (7 %) | 1 (8 %) | 2 (8 %) |
| **Gender n (%)** |  |  |  |
| male | 9 (69 %) | 3 (25 %) | 12 (48 %) |
| female | 4 (31 %) | 9 (75 %) | 13 (52 %) |
| **Highest level of education n (%)** |  |  |  |
| High school | 0 | 0 | 0 |
| Some College | 3 (23 %) | 1 (8 %) | 4 (16 %) |
| Associate degree | 4 (31 %) | 0 | 4 (16 %) |
| Bachelor's degree | 1 (8 %) | 6 (50 %) | 7 (28 %) |
| Master's degree | 2 (15 %) | 1 (8 %) | 3 (12 %) |
| PhD | 3 (23 %) | 4 (33 %) | 7 (28 %) |
| **Race n (%)** |  |  |  |
| White | 10 (77 %) | 8 (67 %) | 18 (72 %) |
| Black/African American | 1 (8 %) | 1 (8 %) | 2 (8 %) |
| Asian | 2 (15 %) | 2 (17 %) | 4 (16 %) |
| Multi-racial | 0 | 1 (8%) | 1 (4%) |
| **Yrs since diagnosis of PD n (%)** |  |  |  |
| ≥ 15 yrs | 1 (8 %) | 3 (25 %) | 4 (16 %) |
| 11 - 14 | 5 (38 %) | 4 (33 %) | 9 (36 %) |
| 6 – 10 | 4 (31 %) | 3 (25 %) | 7 (28 %) |
| 3 – 5 | 3 (23 %) | 2 (17 %) | 5 (20 %) |
| Less than 3 yr | 0 | 0 | 0 |
| **H&Y stage n (%)** |  |  |  |
| Stage 1 | 0 | n/a | 0 |
| Stage 2 | 3 (23 %) | n/a | 3 (23%) |
| Stage 3 | 6 (46 %) | n/a | 6 (46%) |
| Stage 4 | 4 (31 %) | n/a | 4 (31%) |
| **Hospitalization type n (%)** |  |  |  |
| Planned hospitalization | 4 (31 %) | 0 | 4 (16 %) |
| Unplanned hospitalization | 9 (69 %) | 12 (100 %) | 21 (84 %) |

A semi-structured discussion guide, developed de novo by the research team based on a review of the published literature on hospitalizations in PD, was used to structure the focus groups (Appendix 1). As the data collection progressed, the discussion guide was adapted to incorporate new issues raised by the participants. The questions focusing on aspects of hospital admission, inpatient experiences, discharge processes, interactions with the health system and team, and lived experiences of PwPs and CPs thought to be most relevant to patient- and family-centered outcomes. An experienced group moderator (J.S.) used probing questions to further expand the discussion. Prior to the focus groups, all the participants completed a brief questionnaire to capture their demographic information.

All focus groups were conducted virtually on the Zoom platform and were recorded and transcribed verbatim. Each group was 120 minutes in length. Identifiers were stripped from the transcripts, which were reviewed for accuracy. Participants received $20 honoraria. After conducting 7 focus groups, the research team determined that information power was achieved, and recruitment ended.^42^

### Data management and analysis

Transcripts were independently reviewed by a multidisciplinary team of three researchers, including a movement disorders specialist (N.B.), a clinical social worker (J.S.), and a qualitative methods expert (S.G.), to identify emerging concepts related to hospital experiences. During the first phase of analysis, two investigators (N.B. and S.G.) independently read 3 transcripts before convening to define initial topics/concepts and develop a preliminary codebook. Coded data and transcripts were maintained in an electronic database, MAXQDA 2020 (VERBI software, 2019). An inductive thematic approach was used for analysis. The respective coded transcripts were compared during face-to-face meetings (N.B. and S.G.) to assess similarities and discrepancies regarding code names and code application. Based on these consensus meetings, researchers developed a final codebook that was systematically applied to the remaining transcripts. The team continued to review and code transcripts independently, meeting regularly to collaboratively discuss coding decisions and to resolve any coding differences through consensus. All coded transcripts were then reviewed by a third researcher (J.S.) to ensure consistency. ^43^ The varied perspectives of team members yielded a nuanced and robust interpretation of the results and all discrepancies among analysts were resolved. The research team (N.B., J.S., P.M.) analyzed each code and assessed conceptual relationships among them to develop higher-level categories and the relevant sub-themes within each category. The findings were then condensed, and conclusions drawn.

## Results

### Participants

Seven focus groups were conducted. Participants included 12 PwPs (69% in the 65-74 age range and 28% non-white) and 13 CPs, including five PwP-CP dyads, for a total of 25 participants (Table 1). 84% of participants had unplanned hospitalizations and 64% of participants had lived with the PD diagnosis between 6 and 14 years at the time of hospitalization. All CPs reported unplanned hospitalizations of their loved one with PD. Ten participants were hospitalized at academic medical centers. During recruitment, participants were identified by their roles in the healthcare system (patient versus family CP). Initially, the researches planned to create homogenic focus groups that thought would facilitate open discussion. ^45^ However, during the recruitment, several PwPs in more advanced diseases stages expressed a preference for their CPs to be present during focus group and help navigate challenges with speech or slower speed of processing, which made it difficult for them to fully participate in the discussion. In these PwP – CP dyads, the CP commonly participated in discussion by either voicing their own opinion about hospitalization or helping the PwP express their thoughts, thus playing the role of “patient’s voice.” The study included two focus groups consisting solely of CPs, one focus group with only PwPs, and the remaining four focus groups had a mix of participants (Table 2).

**Table 2.** Composition of the focus groups Dyads of PwP – CP are marked with *.

| **Focus group #** | **Subject ID** | **Category of participant** | **Planned or unplanned hospitalization** | **Academic or community hospital admission** | **Gender** | **Race/ Ethnicity** |
| --- | --- | --- | --- | --- | --- | --- |
| 1 | P_1_G_1_CP* | Care partner | Unplanned | Community | F | Multi – racial |
| 1 | P_2_G_1_PwP* | Patient | Unplanned | Community | M | White |
| 1 | P_3_G_1_CP* | Care partner | Unplanned | Community | F | White |
| 1 | P_4_G_1_PwP* | Patient | Unplanned | Community | M | White |
| 1 | P_5_G_1_PwP | Patient | Planned | Academic | F | White |
| 2 | P_6_G_2_PwP | Patient | Unplanned | Community | M | White |
| 2 | P_7_G_2_PwP | Patient | Planned | Academic | M | White |
| 2 | P_8_G_2_PwP* | Patient | Unplanned | Community | F | White |
| 2 | P_9_G_2_CP* | Care partner | Unplanned | Community | M | White |
| 3 | P_10_G_3_CP | Care partner | Unplanned | Academic | F | White |
| 3 | P_11_G_3_CP | Care partner | Unplanned | Community | F | White |
| 3 | P_12_G_3_CP | Care partner | Unplanned | Community | M | Asian |
| 3 | P_13_G_3_CP | Care partner | Unplanned | Community | M | Asian |
| 3 | P_14_G_3_CP | Care partner | Unplanned | Academic | F | White |
| 4 | P_15_G_4_PwP | Patient | Planned | Academic | F | White |
| 4 | P_16_G_4_PwP | Patient | Unplanned | Community | M | Asian |
| 5 | P_17_G_5_CP | Care partner | Unplanned | Academic | F | White |
| 5 | P_18_G_5_CP | Care partner | Unplanned | Academic | F | White |
| 6 | P_19_G_6_PwP | Patient | Unplanned | Academic | M | Asian |
| 6 | P_20_G_6_PwP | Patient | Unplanned | Community | M | African American |
| 6 | P_21_G_6_CP* | Care partner | Unplanned | Community | F | African American |
| 6 | P_22_G_6_PwP* | Patient | Planned | Academic | F | White |
| 7 | P_23_G_7_PwP* | Patient | Unplanned | Academic | M | White |
| 7 | P_24_G_7_CP* | Care partner | Unplanned | Academic | F | White |
| 7 | P_25_G_7_PwP | Patient | Unplanned | Community | M | White |

## Resulting categories

The focus group discussions revealed rich descriptive and thematic data, however, this paper focuses on two specific categories: the impact of the PD diagnosis on the patient and family’s hospital experiences and perceptions of care, and the emergence of distinctive needs of PwPs and CPs during hospitalization.

Tables 3 and 4 present the themes accompanied by the focus group participants’ representative quotes. Each quote is marked with participant number (P#), focus group number (G#), and whether the participant identified as PwP or CP. In the following section, we outline the major themes of each category.

**Table 3.**
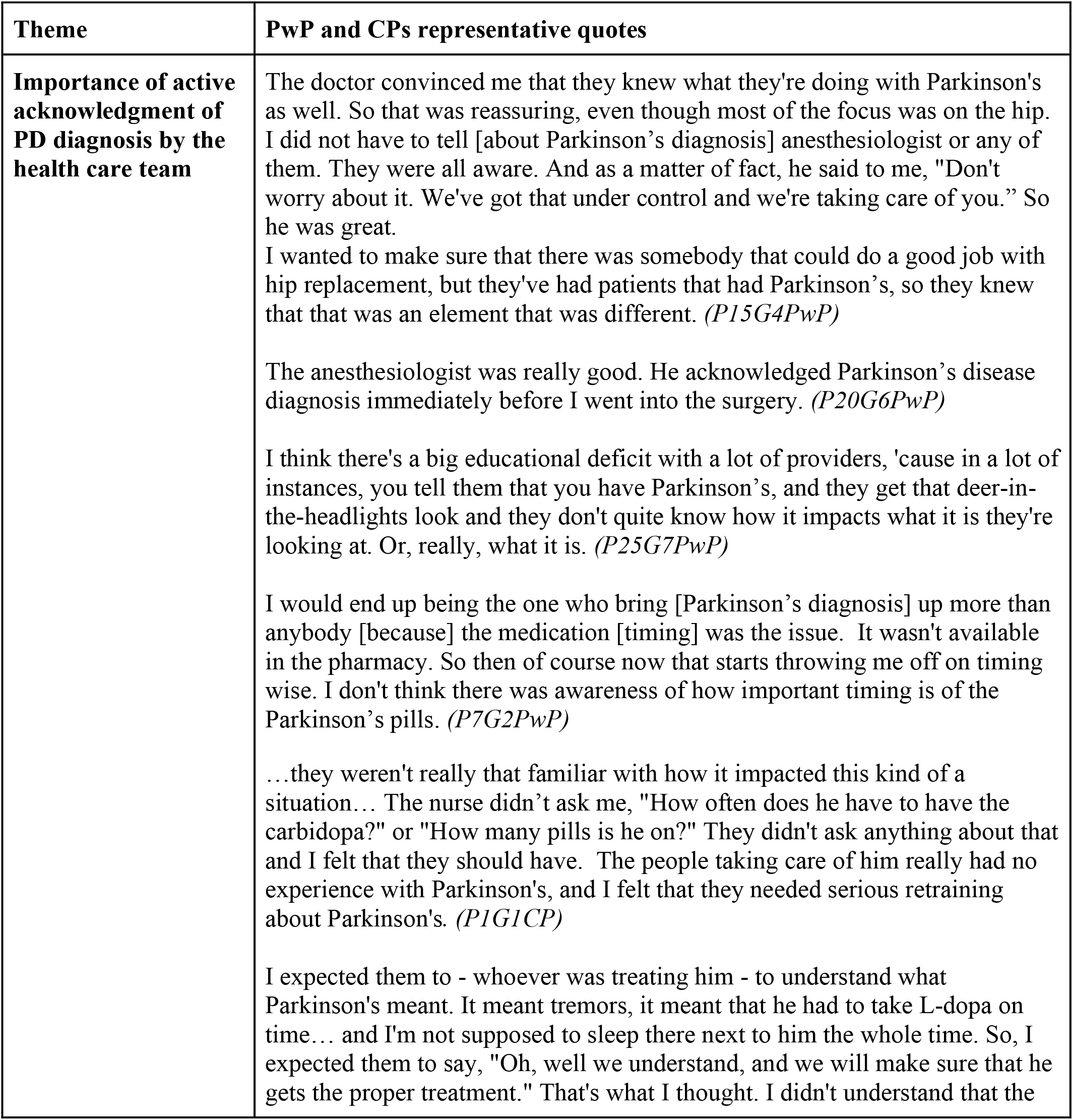
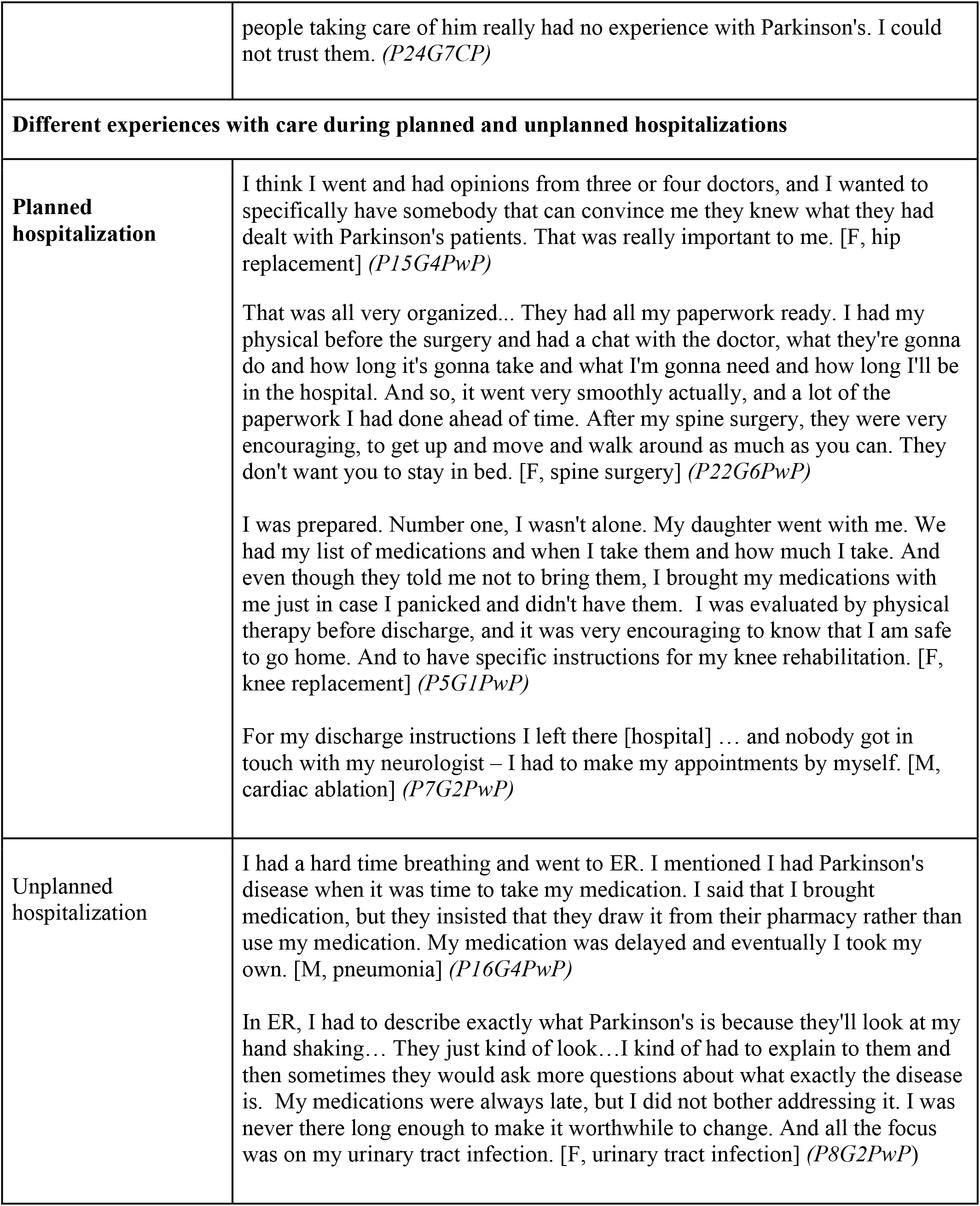
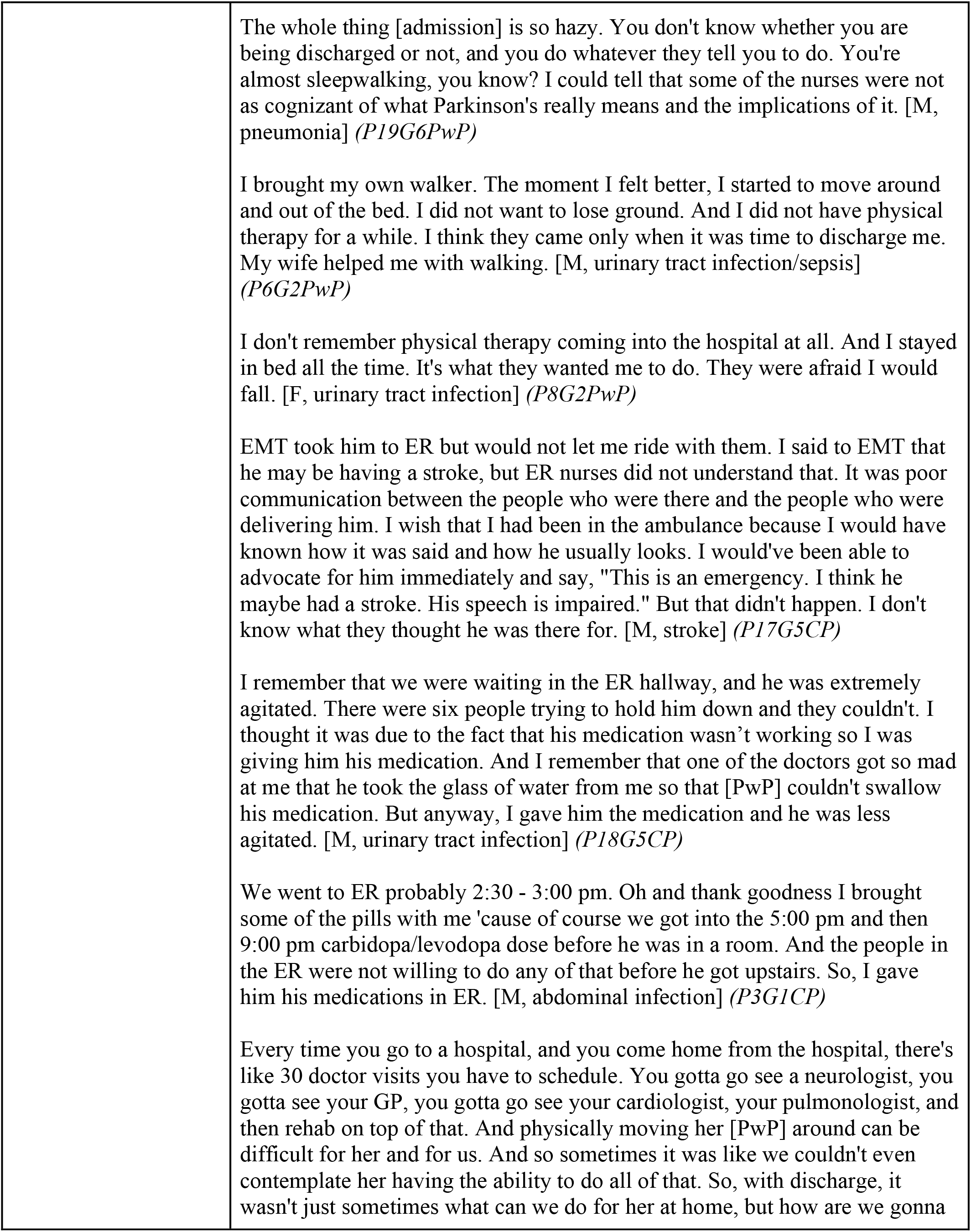
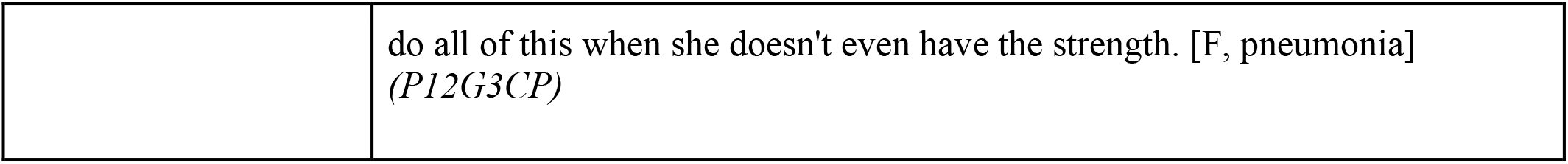
The impact of PD diagnosis on the hospital experience and perception of care among PwP and CPs during hospitalizations with representative quotes. PwP gender and diagnosis of hospitalizations presented in parenthesis.

**Table 4.** The unique needs of PwP and CP during hospitalizations with representative quotes with representative quotes.

| Themes | PwP and CPs representative quotes |
| --- | --- |
| *Dissatisfaction with management of PD medications* |  |
| Inconsistent availability of PD medications | The medication was the issue. I brought my medication with me, they insisted that I couldn't have my own little stash. I needed to go to the pharmacy. And they took down whatever I was supposed to take. So okay, I take the pills that I've got with me, give them to the nurse who puts them down in the pharmacy and then I have to get them brought back up to me. And all of this is taking time that is pushing me off my normal schedule. (P15G4PwP)<br>They said that the hospital didn't have it [carbidopa]. They said, "Well, we'll have to see if the pharmacy can get it." And so I offered to bring it and they didn't want me to bring it. It was after the third day that they finally got it straightened out, so I don't think he had carbidopa until just before time to leave. (P21G6CP) |
| Delays in medication schedule | Every four hours, that has to be taken; otherwise, my reaction is awful. Leg starts kicking, the hands start kicking, the body shakes. So, I mean, it's really an awful feeling. (P19G6PwP)<br>I can tell when I need to take my medications. That's when my tremors start when I'm not on time. I take it three times a day. As it wears off, I take it. I asked for it, it took them an hour to get it. (P8G2PwP)<br>But I would also say when someone with Parkinson's is a patient in the hospital, they are probably under stress due to health issues. They probably need their medication early some of the time, not just on time. (P14G3CP)<br>Our biggest problem was the timing. They could not get the idea that the carbidopa/levodopa had to be given at a certain time particularly in relation to meals. (P3G1CP)<br>We know they're hospital and they're not gonna be on the dime at the exact timing, so you have to be somewhat flexible. But with Parkinson's patients, especially with someone who's taking medication every two hours, more than 15 minutes is very impactful. (P12G3CP) |
| Substitution of medications | They were giving me different looking medications that they assured me were the same thing, just a different manufacturer or whatever it was. Every time they gave me something and I looked at it and questioned it. I was only there for two days so it wasn't an extended time period, but it was very disconcerting for me. (P5G1PwP) |
| Self-administration of medications | Now, what happened was that they didn't have something I needed - I wanna say some ropinirole - they did not have it in stock. So that was a problem. And I quite honestly had my son and my husband go home and they sort of snuck it in... (P15G4PwP)<br>I let my wife know to bring the Neupro patches. It would've been a couple days before it got to the pharmacy, so we just went with mine and I thought that was good 'cause they normally don't allow you to do that. (P25G7PwP)<br>And so I tried to tell them that I spoke to my doctor and that this was all cleared and it was fine, but they were a little hesitant and, at one point, said, "Well, why don't you give us the medication and we'll give it to you?" And I said, "No..." I said, "I wanna keep the medication in my nightstand here so I don't get it mixed up with anything else," and so they finally said that was fine. (P22G6PwP)<br>I wish they were away for patients who are self-aware to be able to be more self-dosing while they're in the hospital, with some limit per day. But they often know their needs better than any staff can. (P16G4PwP)<br>...and so a couple times I gave medication to him when he needed to have it and then I just told them that I'd already given it to him. I know they were not happy but we couldn't wait. It was very hard to get through that this is time sensitive. (P3G1CP)<br>I didn't trust them [nurses] to give it to him, so I wanted to give him his pills. (P24G7CP) |
| ***Desire for flexibility in hospital protocols regarding falls risk and ambulation*** |  |
| limitations in mobility protocols and overall ambulation while in the hospital | It was kind of funny that as soon as I said Parkinson's they put a tag on my hand saying that the fall risk. So after that, they wouldn't let me get off the bed by myself, even though I was pretty able to walk. As long as I'm on the medication, I'm pretty stable. But they put the tag and after that I had to call the nurse every time I want to get off the bed and use the bathroom or anything like that. (P25G7PwP)<br>They had fits because I would get out of bed and I'd have to urinate and they would just go ballistic about me getting out of bed but they wouldn't come right away 'cause they were dealing with other issues so I took it upon myself to. So finally at about 6 in the morning they brought a port-a-potty into the room. But you know they should have done that sooner if they didn't want me getting out of the bed 'cause alarms went off left and right. (P7G2PwP)<br>"You should continue on moving and get out of the bed" that is not something they [healthcare team] discussed at all. (P19G6PwP) |
|  | It was difficult to convince them that I'm pretty capable of getting up and walking by myself. They just wouldn't listen to me. And I didn't make a fight because I knew that they had their protocols and that they're following theirs. And like I said, if my fall risk increases, if I realize that I'm getting weak or I stumble more, I would totally welcome that protocol. I know it is useful. (P16G4PwP)<br>To keep someone in bed even two days, my husband has had to relearn to walk twice during the length of his hospitalizations. And it could be prevented if you were able to get out of bed and walk around the room or down the hall every day and it's just not encouraged. They come and do the physical therapy evaluation before discharge. And they should be getting them out of bed every day. It's so quick that you lose the ability to walk. (P14G3CP)<br>And they wouldn't let him out [to be discharged] because they said his balance was so bad. But he had been in bed for three days. So of course, his balance is bad. That's a long time to be in a hospital where he's totally in bed. He couldn't get up and walk. He just had to be in that bed. (P10G3CP)<br>[ for inpatient PT evaluation] he was able to walk across the room and down the corridor. So there wasn't an issue that time. But then when he eventually started doing it at home in a home setting, it was obvious that maybe it was not possible to do. If he would go outdoors, walk down the driveway, and get into the vehicle or drive somewhere and come back, then he couldn't get out of the vehicle. He would just collapse on the ground.(P1G1CP)<br>For Parkinson's, every patient is different and their needs are different. So when I said, "You know, he needs help getting up out of the bed." And he has to have help getting out of a chair even now, and so I had to tell them that because they would come into the room and say, "Okay, it's time to go to the bathroom," and they would take the pole or let him take the pole. He can't do that, you know? His disability doesn't allow for that...(P24G7CP)<br>Even if not walking but at least just some kind of sitting down exercises, and to take him to have some kind of activities. We really don't have that. It hasn't been happening in a hospital on its own, isn't it? It should happen especially with Parkinson's patients.(P21G6CP) |
| ***The need for disability – informed hospital environment*** |  |
| Preservation of independence in the hospital environment | Sometimes it was difficult, especially the dinner, to cut the food up I mean, 'cause my hands start shaking. (P19G6PwP)<br>My husband would always try to order finger food that he would rather feed himself and he always had difficulty with that hand coordination after surgery |
|  | for several days. And so to think about maybe adding items that are finger foods, don't fall apart when you pick them up. *(P14G3CP)*<br>You don't give a Parkinson's person with tremor a full glass of water because it'll be all over the place. You also offer them a straw and always do a half glass of water. Little things like that make all the difference. *(P24G7CP)* |
| Desire for accommodations for PD – specific care needs | It's hard for me to sleep so when I get to sleep, I'm not really happy when somebody wakes me up and they would come in and have to take my temperature and my blood pressure. *(P7G2PwP)*<br>Every time I've been in the hospital, they keep me fairly close to the nurse's desk and especially during change in shift, you've got all these nurses and everybody congregating at this one spot and they're all talking at once. It gets kind of noisy especially in the middle of the night when they're changing shifts. *(P8G2PwP)*<br>Nurses didn't care that I had Parkinson's and, therefore, disabled in ways that they weren't familiar with. *(P23G7PwP)*<br>It was hard to use urinal. Needless to say, there were times that he would get wet or whatever and I asked [the nurse] if she would help and she said, "No, he needs to learn to just do it himself." And it kind of threw me off 'cause I didn't expect that answer. *(P1G1CP)*<br>The problem is as they get older and sicker, we also get older and sometimes also sicker. It was harder and harder for me to spend those nights in the hospital because those chairs that they give you are very uncomfortable. *(P18G5CP)* |

## 1. Pre – existing PD diagnosis affected participants’ hospital experience and perception of care: “They acknowledged [PD] immediately… that was great!”

### 1.1. Acknowledgment of PD diagnosis by the health care team was important to participants

Although none of the participants’ hospitalizations was directly related to PD symptoms, the presence of a PD diagnosis and whether the health care team (HCT) actively acknowledged the PD diagnosis had a significant impact on the perceptions of care of both for the PwPs and CPs. In both planned and unplanned hospitalizations, trust in the HCT was immediately gained when the team openly acknowledged the patient’s diagnosis of PD and demonstrated knowledge about specific considerations during hospital stays, anesthesia, and post-hospitalization rehabilitation.

### 1.2. Hospitalization experience differed according to whether hospitalizations were planned or unplanned

Overall, the participants’ experiences with planned hospitalization were positive starting from the ability to choose their HCT with previous experience in PD care.

> *“That was really important to me. I wanted to make sure that there was somebody that could do a good job with hip replacement but they’ve had patients that had Parkinson’s, so they knew that that was an element that was different.” (P15G4PwP)*

PwP chose the date of their planned hospitalization to ensure the presence of CP during the hospital stay and after the discharge: *“And so I actually chose that particular surgery time so that I knew my daughter would be around… It was at Christmas time and… She is a teacher, and she was actually off for the next two and a half weeks.” (P5G1PwP)*

During planned hospitalizations, PwPs had support from the rehabilitation services, and were given precise discharge instructions regarding the primary cause of hospitalization. Still, participants admitted to struggling to maintain the timing of their PD medication dosage during the hospital stay and their discharge instructions did not reference their diagnosis of PD.

In contrast, unplanned hospitalizations were described as *“chaotic,”* requiring quick decision-making from either PwPs or CPs on whether an ER visit was warranted. For those with unplanned hospitalizations, not one participant mentioned that they had a plan for contacting their neurologist or primary care physician. PwPs and CPs from multiple focus groups commented on delayed access to PD medications in the ER as well as perceived challenges with care delivery (e.g., HCT ability to perform intravenous cannulation placement or chest X-ray) due to prominent PD symptoms such as tremor. CPs were active participants in the decision to go to the hospital for unplanned hospitalizations, and in some cases, drove PwP to the ER. At the ER and once admitted, CP played an essential role in describing the usual state of health of the PwP and helped communicate any changes in their symptoms from baseline to the HCT.

## 2. The presence of PD-specific care needs posed an additional challenge for participants during hospitalization: “I expect them to be aware of the fact that I’m different.”

Specific needs affecting the experience from admission to discharge were identified, including knowledge of PD medications, proper medication management, improved hospital ambulation protocols, and preservation of independence in the hospital environment.

### 2.1 Numerous hurdles with PD medications management lead to dissatisfaction with hospital care among the participants

Prior to any type of hospitalization, across all focus groups, PwPs and CPs were concerned about the availability of PD medications and, as a result, packed and brought their medications to the hospital. All participants reported issues with consistent and timely delivery of PD medications at all stages of hospitalization, from admission to discharge. For some, medications were substituted or re-arranged without explanation, which created mistrust towards HCT.

> *“They were giving me different looking medications that they assured me was the same thing, just a different manufacturer or whatever it was. Every time they gave me something and I looked at it and questioned it. … it was very disconcerting for me.” (P22G6PwP)*

Most participants reported needing to have continuous discussions about their medication regimens with their care team. Trust in the HCT further eroded when the PwP perceived that the team lacked knowledge of commonly used medications for PD.

> *“Everyone had a general understanding of Parkinson’s, but not what I would consider, really, decent depth. And especially when it came to the medication, that was a tangible way of judging [the team]-that was something that had to happen and they needed to understand why it was important.” (P26G7PwP)*

To ensure the correct medications were taken on time and as prescribed, many PwPs and CPs chose to administer their own medications during hospitalization. This was accomplished with or without nursing staff awareness. One CP explained: *“I didn’t trust them to give it to him, so I wanted to give him his pills.” (P24G7CP)* While acknowledging possible limitations, many participants expressed a desire to see protocols around medication self-administration in the hospital. As one participant shared: *“I wish there was a way for patients who are self-aware to be able to be more self-dosing while they’re in the hospital, with some limit per day. They often know their needs better than any staff can.” (P16G4PwP)*

### 2.2 The restrictive nature of the hospital fall prevention protocols, along with dissuasion of ambulation, was discordant to participants needs to maintain mobility in the hospital

Participants who experienced planned hospitalizations for orthopedic issues received prompt postoperative physical therapy (PT) with encouragement for daily ambulation. However, during unplanned hospitalizations, PwPs struggled to advocate for their ambulation needs and reported limited or no evaluation by PT and decreased mobility due to bed confinement. While CPs were commonly present at the bedsides of PwP, they were unsuccessful in advocating for more physical activity. In all focus groups, both PwPs and CPs remarked that the immobility of the PwP was not a concern for HCT. While some participants actively advocated for more physical activity and an assessment by a PT during their hospital stay, others did not, but still expressed their concerns during their focus group.

There were multiple PwPs with good postural stability and no history of falls who were deemed to be a “fall risk” during their hospitalization. *“It was kind of funny that as soon as I said ‘Parkinson’s’ they put a tag on my hand saying that I have fall risk. So, after that, they wouldn’t let me get off the bed by myself, even though I was able to walk.” (P16G4PwP)* The discrepancy between PwP needs to maintain mobility in the hospital, and the restrictive nature of the hospital fall prevention protocol, along with dissuasion of ambulation, was unsettling to the patients. *“They had me in lockdown mode because I was in the fall risk… I would just attempt to escape from Alcatraz.” (P25G7PwP)* While participants acknowledged fall prevention as an important aspect of hospitalization, not many PwPs mentioned success in their advocacy to the HCT to revert fall prevention protocols despite obvious distress that such protocols created during their hospital stay.

### 2.3 Hospital environment was not accommodating towards participants’ existing motor and non – motor limitations, indicating the need for disability-informed hospital environment

Both CP and PwP participants reported feeling that PwP’s sense of was significantly altered in the hospital. They described the impact of poor fine motor control (due to bradykinesia or tremor) on PwP’s ability to attend to daily tasks, such as eating and preferring finger foods on the menu, drinking from half-filled glasses to prevent spillage, and requiring assistance with managing urinals or pushing buttons on bed controls. Both PwPs and HCT preferred CPs to be at the bedside to aid in communication related to PD (e.g., voice, cognition), although many CPs commented on the lack of accommodations for them, including limited space at the bedside or uncomfortable chairs. Some participants described significantly interrupted night sleep due to vital sign assessments, hearing conversations at the nursing station, or being awakened early to take morning medications. Some CPs observed that sleep interruptions created subsequent confusion and delirium and negatively affected the hospital experience for PwPs. Notwithstanding the reason for hospitalization, when accommodations for PD-specific care needs were included in hospital care, the experience was perceived by PwPs and CPs as more positive than when accommodations were excluded.

## Discussion

Our study used an innovative approach to define care needs of PwPs and CPs in the inpatient setting. By gathering first-hand experiences from direct stakeholders, we used qualitative and patient-centered methods to define the challenges and opportunities for improving hospitalization for PD. Thematic analysis revealed unique needs of PwPs and CPs while in the hospital, including the desire for individualized treatment plans and approaches, and the impact of the PD diagnosis on the perception of care during hospitalization.

Consistent with previous literature, the timely provision of PD medications was a key factor in the experience of and satisfaction with care for participants. ^37,46,47^ In a systematic review examining the prevalence of adverse events related to medication errors, 31% of PwPs expressed dissatisfaction in the way their PD was managed. ^47^A more recent study focusing on motor outcomes identified medication errors as the most important factor in motor deterioration during hospitalization. ^14^ Owing to challenges with medications in the hospital, most participants in our study proceeded with or desired medication self–management. Studies in other patient populations demonstrated the benefits of carefully applying validated medication self– administration protocols during hospitalization and after discharge. ^48–50^ The potential benefits and barriers to PD medication self-administration have been explored in outpatient settings; however, no study to date has assessed attitudes toward inpatient medication self-management in the PD population.^51,52^ Strategic and evidence-based medication self-management protocols for PwPs in the early stages or with support of CPs could empower PwPs and CPs and alleviate the workload on hospital staff.

Participants highlighted an important opportunity to improve PCC through individualized assessment of fall risk and flexibility in fall prevention protocols. To our knowledge, the study of falls and fall prevention protocols in hospitalized PwPs does not exist, even though gait and balance deficits were found in 41% of hospitalized PD patients, and prospective studies documented falls in up to 70% of PwPs. ^53,54^ In older adults, a multidisciplinary and patient-centered approach to the development and implementation of hospital fall prevention protocols has been beneficial and could serve as a roadmap for similar quality improvement initiatives for PwPs. ^55–57^ Participants in our study also strongly advocated for safe mobilization and early assessment by rehabilitation therapists during their hospital stay because of their fear or the reality of worsening PD motor symptoms due to immobility. Although there is a lack of literature on the safety and feasibility of early mobilization for hospitalized PwPs on general wards, studies show the benefits of early mobilization after surgery in PD. ^58,59^ Walking during hospitalization is effective for older adults, promoting mobility, shortening hospital stays, and increasing likelihood of discharge to home. ^60^ Interventions to encourage mobility in this population show promise in preventing hospital-associated functional decline and maintaining prehospitalization mobility. ^61–63^ Reported barriers to physical activity during hospitalization include insufficient staffing to assist with or encourage mobility, illness symptoms, fear of falls, and a discouraging hospital environment. Our study participants alluded to similar barriers to mobilization during their hospital stays. ^64–66^ In addition to further research on fall prevention protocols for hospitalized PwPs, identifying patients with low fall risk and encouraging safe ambulation could be the first step to translate the well-established benefits of sustained mobility from outpatient to inpatient care for PD and to empower PwPs and CPs during hospitalization.^67^

In our study, nearly two-thirds of participants lived with the PD diagnosis for more than 6 years, and 84% experienced unplanned hospitalizations, emphasizing the complexity of care in the mid- and later stages of PD. The participants’ descriptions of challenges with navigating the hospital environment, including but not limited to tremor preventing the ease of intravenous cannulation placement, difficulty picking up and swallowing food that was served, and using hospital equipment like nurse call buttons, were not anticipated by the researchers when this study was designed. These PD-related challenges point to the hidden impact of hospitalization on one’s sense of independence. In addition, many CPs mentioned worsening of cognitive function or the development of delirium in PwPs while hospitalized. Our study methods precluded us from identifying specific practices implemented for delirium prevention; however, participants in multiple focus groups mentioned poor sleep protection for PwP during hospitalization. This was similar for CPs, who left the hospital feeling exhausted from reportedly sitting in uncomfortable chairs, monitoring and speaking for their PwP, and continuing to care for their partners once discharged home. Patients diagnosed with PD are fivefold more likely to be treated for delirium than patients from the general population, which may be related to non-motor symptoms in PD, such as dementia, cognitive impairment, and sleep disturbances. ^8,68,69^ Since hospitalization places older adults and PwPs alike at risk for new or worsening disability and reduces likelihood of recovery, several successful interventions have been employed to modify hospital environment and improve patient experience and outcomes. ^57,61,62,70^ The hospital environment has a significant impact on patient satisfaction with care, and thus, it could be beneficial to develop and adopt customized hospital accommodations for PwPs to optimize outcomes and decrease risk of complications. ^23,71^

When patients with chronic conditions are admitted to the hospital, they are expected to switch from being the leader of their own care to being a passive consumer who resumes self-management only upon discharge. Consequently, during hospitalization, the combined stress of acute and chronic illness, set against the background of ongoing pressure to advocate for their unique needs, may be all-consuming for PwPs and CPs. Yet, this can be easily overlooked by HCTs as they are focused on medical management of the acute condition that caused hospitalization. In our study, PwPs and CPs sought active acknowledgement of PD diagnosis by their HCT and adjustment of the hospital communications, protocols or even environment, all of which underscore the impact of PD diagnosis on their perceptions of and experiences with inpatient care. Chronic care advocates argue that hospitals will continue to play a key role in chronic disease care, despite how many acute hospitalizations can be avoided, as most chronic conditions are characterized by acute exacerbations requiring admission. ^72,73^ Innovative care delivery models, such as the Chronic Care Model, recognize the importance of better preparing hospitals for a role in chronic illness management and demonstrate positive outcomes associated with specialized knowledge of PD among inpatient HCTs. ^74,75^ Thus, key findings from our study support acknowledging and accommodating the intersectional needs between the chronic condition of PD and the acute reason for hospitalization of the PwP.

The strength of our study is the use of purposeful sampling, a technique widely used in qualitative research, to identify and select information-rich cases for the most effective use of limited resources. ^42^ Purposive sampling allowed us to identify and select individuals in different stages of PD and ensure that we would capture maximum variation of hospitalization experiences. Qualitative analysis can reveal themes in the data that otherwise may be difficult to identify using quantitative approaches. Focus groups, as a qualitative method, carried an additional strength by creating information – rich data. Focus groups allowed people to discuss the relevant topics with other PwPs and CPs using their own language, to build upon each other’s accounts and promoted ’memory synergy’, bringing forth a ’collective memory’ of varied perspectives on similar experiences during hospitalization. ^76^ One of the limitations of our study is that we were unable to recruit CPs who experienced planned hospitalizations with their PwP, and, as a result, this perspective was not represented in our focus groups. Our sample was largely white, despite having intentionally expanded our recruitment efforts to include PwPs and CPs from diverse demographic backgrounds. Racial and ethnic differences in diagnosis, care experiences, and treatment utilization with PD are well known.^77^ Therefore, the findings from this study likely cannot be generalized to the overall PD population and must be further validated in people with varied racial, ethnic, socioeconomic, and clinical backgrounds. Because the focus groups occurred months after their hospital stays, participants’ reports were subject to recall bias, and their nonclinical knowledge may have restricted their abilities to identify all factors impacting their hospitalizations. Despite these limitations, this study provided a novel opportunity for PwPs and CPs to describe their own realities of their hospitalization experiences.

Our study adds to the canon of literature on hospital care for PD. Still, several concepts brought forth by this study warrant further exploration. There is an opportunity to further investigate the role and impact of advocacy by PwPs and CPs on healthcare delivery, as well as explore methodology to capture the real-time experiences of PwPs and CPs during hospitalization, as has been accomplished in other medical conditions. ^78^ Additionally, the methods and findings of this study serve as good starting points for understanding the hospital experiences of those with atypical parkinsonian syndromes, including progressive supranuclear palsy and multiple system atrophy, given the complexity of symptoms, rapid disease progression, profound lack of awareness of these rarer neurodegenerative diagnoses within the medical community, and the current dearth of research on hospital care for atypical parkinsonism. ^79,80^

## Conclusion

Our qualitative study draws attention to the significant impact a PD diagnosis can have on planned and unplanned hospital stays, even when the reason for care is not directly related to PD. It highlights the plethora of unique needs PwPs and their CPs have during hospitalization. Findings from this study can be used to inform patient-centered interventions aimed at improving the experience with hospital care for PD, including tools that help PwPs prepare for and advocate during hospitalization as well as ensuring flexibility, as appropriate, within hospital protocols. Empowering PwPs and CPs to communicate their questions, concerns, goals, and needs, both generally and regarding PD, with HCT in the hospital setting, thus applying the principles of PCC, could lead to the care they desire and set them up for higher likelihood of positive outcomes following hospitalization.

## Data Availability

All data produced in the present study are available upon reasonable request to the authors

## Acknowledgement

We extend our deepest appreciation to each and every study participant. Their altruistic contribution and willingness to share their experiences has not only enriched our lives, but also have been invaluable in advancing our understanding of PD care during hospitalization and ultimately enhancing the lives of the entire Parkinson disease community.

## Appendix 1. Focus group discussion guide

Appendix 1. Focus group discussion guide.
| Topics | Questions | Prompts | Probes |
| --- | --- | --- | --- |
| **Before hospitalization** |  |  |  |
| Introduction | Please introduce yourself | How long ago was your hospitalization? |  |
| Before Hospital | What made you decide to go to the hospital? | Was it a planned visit? (e.g., scheduled surgery)<br>Was it an unexpected situation? (e.g., change in functioning, fall) | Before coming to the hospital (either for planned or unplanned admission), how did you prepare for the hospitalization?<br>Did you call you neurologist and they suggested you go? PCP? Urgent care? Called 911? |
| **In the Hospital** |  |  |  |
| Patient and team interaction | What is one thing you feel went well during your experience? | Can you tell me more about that? |  |
|  | To what degree do you feel that the medical team met your needs? | Can you tell me more about that? |  |
|  | How do you feel your diagnosis of Parkinson's disease impacted how you were treated, if at all? | Were you treated differently because of Parkinson's? In what ways? | Has anybody else had a similar experience?<br>Did anybody have a different experience? |
|  | What was your impression of how knowledgeable your treatment team was of Parkinson's disease? | Do you have an example? | Has anybody else had a similar experience?<br>Did anybody have a different experience? |
|  | To what extent were you involved in medical/care decisions? |  | Does anybody else have anything different to add? |
|  | To what extent was your family member or care provider involved in medical/care decisions? | Did medical providers involve both you and your family member/care<br>provider in decisions? Please elaborate.<br>Did medical providers communicate differently or the same with you as they did with your family member/care provider? Please elaborate. |  |
| Medications and PD | While in the hospital, were you given your Parkinson's medications as prescribed, on time? | If no, please elaborate. | Did you need to advocate for yourself/partner to get your medications on time? |
|  | While in the hospital, were your Parkinson's medications substituted or adjusted by the team? | If yes, please elaborate. | Did you need to advocate for yourself/partner to get your medications on time?<br>Did you need to bring your own medications to use in the hospital?<br>Did you use your own medications? |
| Rehab service and PD | Did you need rehabilitation or PT/OT evaluation while in the hospital? | If yes, was it provided? How soon? | If you are using cane or a walker on regular basis at home, were those provided for you in the hospital?<br>Were you encouraged to ambulate in the hospital? |
| Hospital environment | How did you find your environment in the hospital? | Were you able to attend to your activities (bathroom, grooming,<br>toileting)? Did you needed help?<br>Do you think any of your PD symptoms were impacted by hospital environment? | If you needed help, who provided it? |
| **Advocacy and empowerment in the hospital** |  |  |  |
| Aware in Care kit | Do you have an Aware in Care kit and if so, did you bring it with you? | If yes, how were you able to utilize the contents during the hospitalization? |  |
| Advance care directives | Have you had any advance care directive paperwork prior to hospitalization? | Have you brought any advance care paperwork in?<br>Who, if anybody, reviewed it? |  |
| Advice for future hospitalization/ to other patients | Thinking on your experiences that you shared today, are there ways your hospitalization, in regards to your Parkinson's care, could have been made better? | What advice would you give someone with Parkinson's who needs to be hospitalized? |  |
|  | Anything else that comes to mind that we did not talked about in our discussion? | Tell me more about that |  |

